# Syndromic surveillance as a predictive tool for health-related school absences in COVID-19 Sentinel Schools in Catalonia, Spain

**DOI:** 10.1101/2023.03.24.23287681

**Authors:** Fabiana Ganem, Lucia Alonso, Andreu Colom-Cadena, Anna Bordas, Cinta Folch, Antoni Soriano-Arandes, Jordi Casabona, COVID-19 Sentinel School Network Study Group of Catalonia

## Abstract

Monitoring influenza-like illness through syndromic surveillance could be an important strategy in the COVID-19 emergence scenario. The study aims to implement syndromic surveillance for children aged 6-11 years in COVID-19 sentinel schools in Catalonia. Data collection was made by self-applied survey to collect daily health status and symptoms. We proceed logistic mixed models and a Latent Class Analysis to investigate associations with syndromes and school absence. Were enrolled 135 students (2163 person-days) that filled 1536 surveys and 60 participants reported illness (29.52 by 100 person/day) and registered 189 absence events, 62 of them (32.8%) related to health reasons. Subgroups of influenza-like illness were founded such as a significantly and positively association with school absences. The findings of this study can be applied to the detection of health events, and association with school absences, offering an opportunity for quick action, or simply for monitoring and understanding the students’ health situation.

**ARTICLE SUMMARY LINE:** This study confirms the relevance of syndromic surveillance in students from 6 to 11 years of age as a strategy to timely detect events that can cause school absence, either to support public health actions by applying analytical models that improve their potential in providing systematized information, or to monitor and understand the health situation of students, thus offering an opportunity for rapid action.

## Introduction

Syndromic surveillance referees a complementary strategy to traditional forms of public health surveillance that aimed early detection of health events [1], faster than routine surveillance, through sensitive data supporting more effective public health actions [2–6]. This approach can have different methodologies, such as monitoring data routinely collected from ambulatorial or primary care, or, through specific systems designed to this purpose [7,8].

The main advantage of a syndromic approach is that there is no need to use data validated by medical professionals and/or laboratory diagnoses, instead, available health information is collected in near real time and can be used to report trends as quickly as possible (Lai et al. 2021). Moreover, many of data collected utilize voluntary reports and depends on human resources and an infrastructure to provide real-time analysis [5,9].

The COVID-19 pandemic brought up several challenges to the public health surveillance, regarding data collection and interpretation until changes in the health system structure, imposing to the traditional surveillance new and adapted approaches to the currently situation [10]. Thus, syndromic surveillance can use no-conventional indicators orientated to early detection [11]. Besides COVID-19, upper respiratory infections (URI) caused by several viruses such as rhinovirus, coronavirus, respiratory syncytial virus, adenovirus and influenza, have a high prevalence, especially in children [12], being a common cause for school absence [13], and responsible for most of the healthcare visits during epidemic seasons [14].

Younger children do not have the skills to report in detail their health status, or to express themselves the potential impact on their daily routine, especially regarding clinical aspects [14]. Therefore, research of tools developed and adapted for children can serve as a guide for capturing information about mild symptoms in a population that not always seek health-care attention at the primary health level [6].

The Catalonia COVID-19 Sentinel Schools Network (CSSNC) is a project that aims to monitor and evaluate the course and effect of the COVID-19 pandemic among students, parents and staff of 23 Catalan sentinel schools. The project was consolidated through several studies that evaluated the incidence and prevalence of SARS-CoV-2 and its association with multilevel determinants, in addition to the development of associated pilot projects to validate complementary surveillance strategies aimed at this population, such as CO2 monitoring, feasibility of public health measures and sentinel surveillance based on school absence caused by respiratory viruses. The study protocol and early results were previously published by (Bordas et al., 2022 and Ganem et al., 2022).

The planning and implementation of complementary surveillance depend on the event to be monitored, population and data available. In this case, to investigate the better approaches among sentinel school population we developed a pilot study to initially answer three questions (i) which symptoms and health conditions are most associated with school absences? (ii) the symptoms severity are the good predictors for school absence? and (iii) which analysis model can capture health events in the daily monitoring of the school sentinel population?

The objective of this study was to implement a methodology for syndromic surveillance for children aged 6-11 years and verify its application as a predictive tool for the health events and health-related school absence in a setting of COVID-19 sentinel schools in Catalonia (CSSNC), Spain during the 2021-2022 academic year.

## Materials and methods

### Study design and population

This is a pilot study that applied a methodology to collect data on symptomatology and school absences among elementary school children from schools participating in the CSSNC. The intervention initially involved the following steps: (i) review of validated instruments to collect respiratory symptoms and school absences adapted for children [6,11,12,14], (ii) development of a short instrument for data collection among study participants, (iii) selection of schools participating in the study, (iv) application of the research in selected schools and (v) data analysis.

The study population included 135 students aged between 6 and 11 years from three primary schools in Catalonia between 16 May and 3 June 2022, spring of 2021-2022 academic year.

### Data collection, variables and outcomes

Initially, we contacted the school staff enrolled in the CSSNC to present the project, inform the procedures and invite them to participate in this specific study. Once the intention to participate was expressed, the schools were included, and a meeting was held to explain the logistics of the intervention and to select the participating classes. The schools participating in the pilot received a package containing the questionnaires and were responsible for distributing them daily to students who answered anonymously. The tutor of each class was responsible for sealing the envelopes containing the completed questionnaires, identifying them only with the name of the school, which were opened by the project members to enter the data in the REDCap project database. All data collected were anonymized.

Data collection was carried out using two short questionnaires only in paper version. The first questionnaire, addressed for the children, contained two questions about the individual daily health status, (i) *How do you feel today*, in which the student could answer the following options: very well, a little sick, sick and very sick; and (ii) *Do you have any of these symptoms*, presented the student a list of respiratory symptoms (fever, sore throat, tiredness, runny nose, cough, sneezing and abdominal pain) on a 4-point severity scale whose options were: I don’t have, a little, quite a lot, a lot. Lastly, we asked if he/she *was able to answer on his/her own or needed help from someone*, because this questionnaire could be filled in two different ways, with the support of the teachers/classmates and by themself. The second questionnaire was filled by the teacher to collect the number of school absences, classifying them by health reasons, by reasons not related to health and by unknown reasons. The variables included in this study were presented in the table 1.

**Table 1.** Summary of variables included in the study, CSSNC, Catalonia, Spain

| **Question/variable** | **Coding** |
| --- | --- |
| *How do you feel today?*<sup>1</sup> | Very well, a little sick, sick and very sick |
| Symptoms available in the questionnaire | Fever, sore throat, tiredness, runny nose, cough, sneezing and abdominal pain |
| *How severe are your symptoms?*<sup>2</sup> | I don't have, a little, quite a lot, a lot (no, mild, moderate and severe) |
| *Answered on his own or needed help?*<sup>3</sup> | By myself, with some help |
| Number of participants | N |
| Age range | Assumed according to the student's school cycle |
| Person at risk | Calculated in this study |
| Number of surveys applied | N |
| Number of surveys completed | By themselves or with help |
| Self-reported health status | Very well and illness (little sick, sick or very sick) |
| Number of participants that reported illness | N, frequency, stratified by severity and by incidence rate |
| Incidence rate of self-reported illness | By 100 person/day |
| Cumulated incidence of self-reported illness | By 100 person/day |
| Number of absences | By health reasons, other reasons and unknown reasons |
| LCA subgroups | Symptoms cluster |

Original version in Catalan 1. Com et sents avui? molt bé, una mica malalt, malalt, molt malalt; 2. Tens algun d'aquests símptomes? No tinc, Una mica, Bastant, Molt and 3. He completat aquesta pàgina: Jo sol, amb una mica d'ajuda d'un company/a o mestre

We defined a school absence as *the daily absence of a student at the school center*. To investigate the association with symptoms, we considered that one student had an absence when he/she had not filled the questionnaire corresponding to that specific day. To validate this assumption, we calculated the total number of missing questionnaires that day and compared to the number of absences reported by the teacher, not considering school trips day.

### Data analysis

We are presenting a descriptive summary of the study population characteristics, and the frequency of the outcomes. We performed a Latent Class Analysis (LCA) to describe potential groups of students with similar symptomatology. LCA consists in categorize latent subgroups, proceeding a cluster analysis based on multivariate binary observations, to reduce the number of variables [17]. First, to remove the random effect of students, we decided to aggregate the repeated measures for each participant into only one. In particular, we considered two approaches: i) to aggregate the symptoms by each week independently, that is, to have one measure for each individual which indicated whether they had each symptom one particular week, ii) to aggregate the symptoms at all study period. We choose the models with 2 and 3 latent classes since models 4 and 5 returned classes with small N and without epidemiological coherence. All these analyses were adjusted by school.

For each symptom and self-reported health status (“how do you feel today”) for those who informed “a little sick”, “sick” or “very sick” was calculated the incidence rate (IR) per 100 person/day that is the number of a new reports (I) per unit of person-time at risk and the cumulative incidence, the proportion of new reports among an initially disease-free population, for the period of the study, using the following equations.

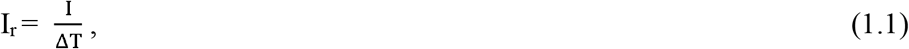

where Δ*T* is the total time under risk of the study population

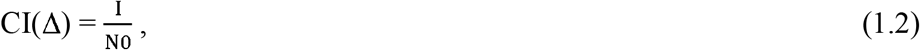

where *N*_*0*_ is the size of the initially disease-free population and *I* the number of new reports (incident) during the study period.

For the univariate analysis, we proceeded a logistic mixed model with subject and school identifiers as random intercepts. First, we investigated the association between one symptom and absence in the next school day, then, we investigated the association between the severity of a symptom and absence. Finally, we performed a multivariate analysis through a forward stepwise regression, but we only obtained one significant variable after selection, so we decided not to include it.

All statistical analyses had been performed in R (version 4.1.2).

## Ethics approval

This study was approved on 17 December 2020 by the Ethical Committee of the Foundation University Institute for Research in Primary Health Care Jordi Gol i Gurina (IDIAPJGol) (code 20/192-PCV). Informed consent providing information about anonymity, confidentiality, use of the collected data, risks, and general information about the study, was signed for school staff, parents for those children under 16 years and student with 16 years or older. Participants were free to decline/withdraw consent at any time without providing a reason and without being subject to any resulting detriment.

## Results

Regarding participation and data collected, there were enrolled in this study 135 students, representing 2163 person-days. We obtained 1536 completed surveys, 1356 (88.2%) filled by themselves (without any help). During the study period, 60 participants reported illness at least one time. The IR of new self-reported illness was 2.32 by 100 person/day, and the cumulative incidence of self-reported illness was 29.52 by 100 person/day. The 135 children generated 189 absence events during the study period, 62 (32.8%) of them related to health reasons (Table 2).

**Table 2.** Summary of participation and descriptive results of the syndromic surveillance pilot study, CSSNC, Catalonia, Spain, May-June 2022

| **Summary of participation and results** | **n** | **%** |
| --- | --- | --- |
| Number of participants enrolled in this study | 135 | 100 |
| Age range | 6-11 | - |
| Person-days<sup>1</sup> | 2163 | - |
| Number of surveys applied | 1601 | 100 |
| Number of surveys completed | 1536 | 95.9 |
| By themselves | 1356 | 88.3 |
| With help | 180 | 11.7 |
| Number of participants that reported feeling very well | 128 | - |
| Number of participants that reported illness (little sick, sick or very sick) | 60 | - |
| Incidence rate of self-reported illness (by 100 person/day)<sup>2</sup> | 2.32 | - |
| Cumulated incidence of self-reported illness<sup>3</sup> | 29.52 | - |
| Number of absences registered | 189 | - |
| For health reasons | 62 | 32.8 |
| For other reasons | 46 | 24.3 |
| For unknown reasons | 81 | 42.9 |

<sup>1</sup> Sum of the time-at-risk for participants overall, covering the entire study period, including weekends.
<sup>2</sup> Incidence rate per 100 person/day is the number of a new reports per unit of person-time at risk.
<sup>3</sup> Cumulative incidence is the proportion of new reports among an initially disease-free population.

The self-reported health status and symptoms were summarized in Table 3. The frequency of students that reported at least one time being sick was 44.44%, during the study period, and the overall the severity average was 1.22 in a 4-point scale. Regarding the self-reported symptoms, runny nose (75.56%), tiredness (72.59%) and cough (61.48%) were the most frequent (table 3).

**Table 3.**
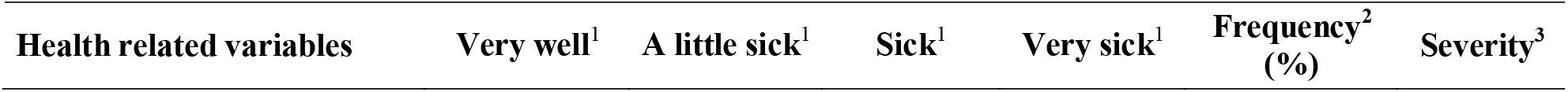
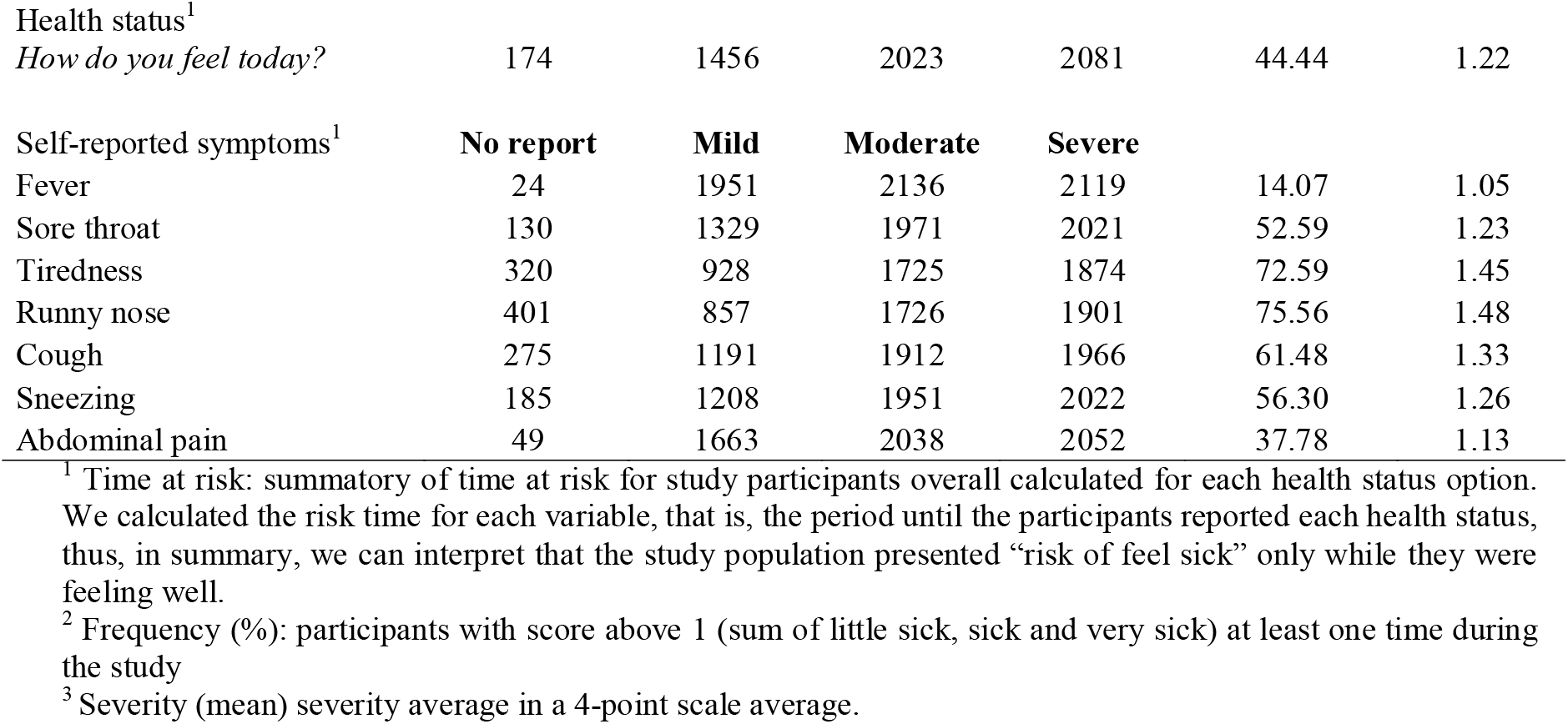
Occurrence, frequency and average severity of self-reported symptoms among study participants, CSSNC, Catalonia, Spain. May-June 2022.

| **Health related variables** | **Very well<sup>1</sup>** | **A little sick<sup>1</sup>** | **Sick<sup>1</sup>** | **Very sick<sup>1</sup>** | **Frequency<sup>2</sup>**<br>**(%)** | **Severity<sup>3</sup>** |
| --- | --- | --- | --- | --- | --- | --- |
| Health status<sup>1</sup> |  |  |  |  |  |  |
| *How do you feel today?* | 174 | 1456 | 2023 | 2081 | 44.44 | 1.22 |
| Self-reported symptoms<sup>1</sup> | **No report** | **Mild** | **Moderate** | **Severe** |  |  |
| Fever | 24 | 1951 | 2136 | 2119 | 14.07 | 1.05 |
| Sore throat | 130 | 1329 | 1971 | 2021 | 52.59 | 1.23 |
| Tiredness | 320 | 928 | 1725 | 1874 | 72.59 | 1.45 |
| Runny nose | 401 | 857 | 1726 | 1901 | 75.56 | 1.48 |
| Cough | 275 | 1191 | 1912 | 1966 | 61.48 | 1.33 |
| Sneezing | 185 | 1208 | 1951 | 2022 | 56.30 | 1.26 |
| Abdominal pain | 49 | 1663 | 2038 | 2052 | 37.78 | 1.13 |

<sup>1</sup> Time at risk: summatory of time at risk for study participants overall calculated for each health status option. We calculated the risk time for each variable, that is, the period until the participants reported each health status, thus, in summary, we can interpret that the study population presented “risk of feel sick” only while they were feeling well.
<sup>2</sup> Frequency (%): participants with score above 1 (sum of little sick, sick and very sick) at least one time during the study
<sup>3</sup> Severity (mean) severity average in a 4-point scale average.

### Latent Class Model

We proceeded to two models with 2 (fig 1a) and 3 (fig 1b) latent classes to describe groups of frequent symptoms reported by the student at least once during the study period. In both models, at least one cluster was formed with a set of symptoms that resembles ILI. In the model with 2 latent classes, there is more variability between the clusters, that is, one group with few symptoms self-reported and a group with many ILI symptoms (2). With 3 latent classes, we can observe a group of ILI symptoms (1), an intermediary group (2) without sore throat and abdominal pain, and a group with few self-reported symptoms (3), (Fig 1a and 1b).

**Fig 1.**
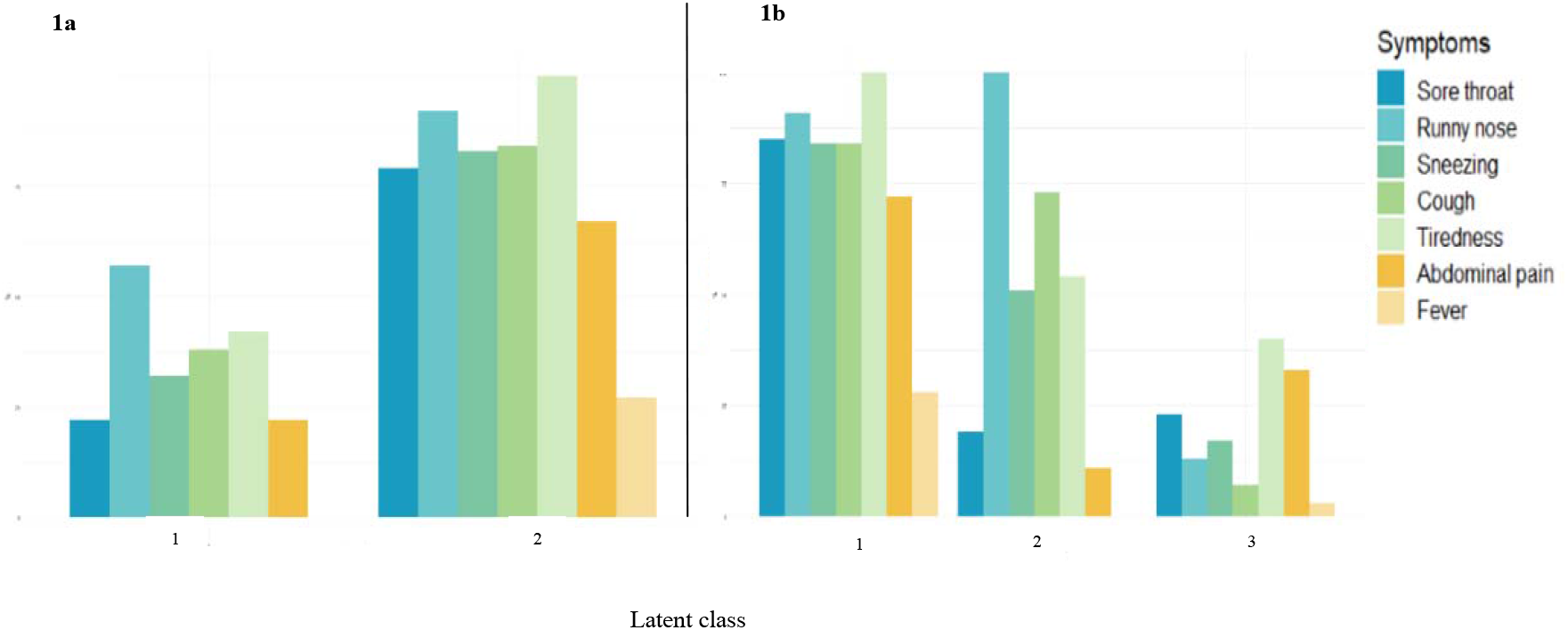
Graphic representation of the syndromes clusters through LCA model with (1a) two latent classes and (1b) three latent classes.

We also built the same LCA analysis through the study weeks to describe variations in clustering symptoms during the study period (Fig 2a and 2b).

**Fig 2.**
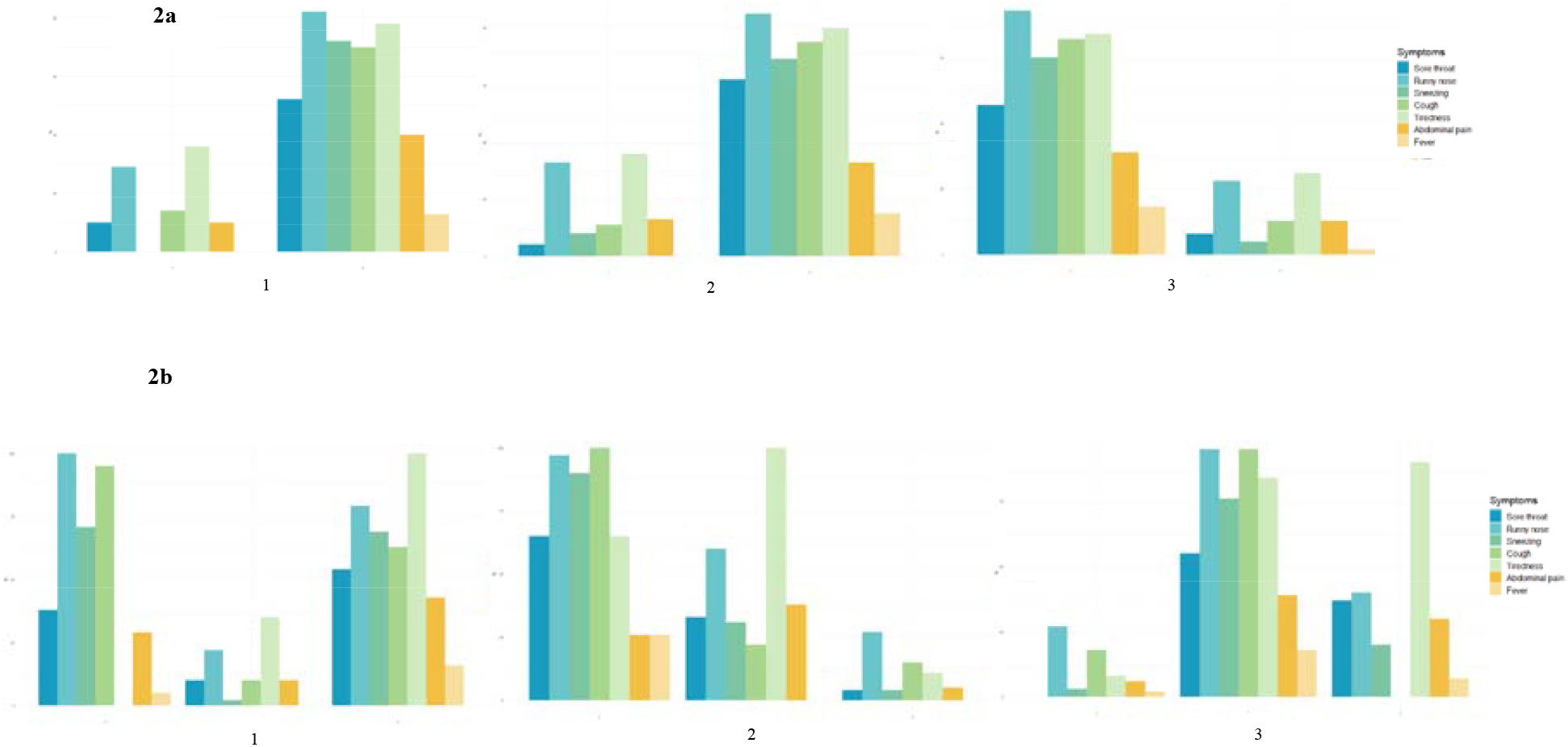
Weekly graphic representing changes in the syndromic cluster through LCA model with (2a) two latent classes and (2b) three latent classes.

### Association between self-reported daily health status, symptoms and school absence

The association between self-reported illness overall or stratified by severity and absence was not significant. We found a significant and positive association between absence and sore throat (OR 2.23), mild sore throat (OR 2.86); moderate cough (OR 4.00); sneezing (2.37), moderate sneezing (OR 6,06), severe sneezing (OR 4,06); and moderate abdominal pain (OR 4.00) (Table 4).

**Table 4.** Association between perceived daily health status, symptoms and school absence among students participating in the Sentinel School syndromic surveillance pilot study, CSSNC, Catalonia, Spain, May-June 2022.

| Variables | Incidence<sup>2</sup><br>(100<br>person/day) | CI95% | Accumulated<br>incidence<sup>3</sup> | CI95% | Univariate<sup>4</sup> |  |  |
| --- | --- | --- | --- | --- | --- | --- | --- |
|  |  |  |  |  | OR | CI95% | p-value |
| Self-reported illness | 2.32 | 1.50-3.14 | 29.52 | 20.80-38.25 | 1,36 | 0.74-2.46 | 0.321 |
| How do you feel today? |  |  |  |  |  |  |  |
| A little sick |  |  |  |  | 1,13 | 0.55-2.27 | 0.744 |
| Sick |  |  |  |  | 2,79 | 0.86-8.99 | 0.085 |
| Very sick |  |  |  |  | 1,44 | 0.30-6.78 | 0.648 |
| **Symptoms** |  |  |  |  |  |  |  |
| Fever<sup>1</sup> | 0.66 | 0.31-1.03 | 10.15 | 4.92-15.39 | 0,65 | 0.15-2.74 | 0.553 |
| Mild | - | - | - | - | - | - | - |
| Moderate | - | - | - | - | - | - | - |
| Severe | - | - | - | - | - | - | - |
| Sore throat | 3.20 | 2.21-4.19 | 38.83 | 29.42-48.25 | 2,11 | 1.18-3.77 | 0.011 |
| Mild | - | - | - | - | 2,86 | 1.51-5.40 | 0.001 |
| Moderate and severe | - | - | - | - | 1,00 | 0.34-2.91 | 0.993 |
| Runny nose | 4.97 | 3.35-6.61 | 52.94 | 41.08-64.80 | 1,26 | 0.77-2.04 | 0.351 |
| Mild | - | - | - | - | 1,01 | 0.58-1.76 | 0.970 |
| Moderate | - | - | - | - | 2,02 | 0.67-6.04 | 0.210 |
| Severe | - | - | - | - | 2,44 | 0.95-6.21 | 0.061 |
| Cough | 3.11 | 2.03-4.19 | 38.55 | 28.08-49.03 | 1,36 | 0.80-2.31 | 0.253 |
| Mild | - | - | - | - | 0,76 | 0.36-1.56 | 0.449 |
| Moderate | - | - | - | - | 4,00 | 1.67-9.56 | 0.002 |
| Severe | - | - | - | - | 2,07 | 0.69-6.11 | 0.190 |
| Sneezing | 2.80 | 1.80-3.81 | 34.09 | 24.19-43.99 | 2,25 | 1.32-3.83 | 0.003 |
| Mild | - | - | - | - | 1,66 | 0.89-3.09 | 0.108 |
| Moderate | - | - | - | - | 6,06 | 2.00-18.25 | 0.001 |
| Severe | - | - | - | - | 4,06 | 1.30-12.66 | 0.016 |
| Abdominal pain | 1.96 | 1.26-2.67 | 26.54 | 18.41-34.69 | 1,44 | 0.70-2.95 | 0.318 |
| Mild | - | - | - | - | 0,84 | 0.30-2.34 | 0.737 |
| Moderate | - | - | - | - | 4,00 | 1.15-13.88 | 0.029 |
| Severe | - | - | - | - | 1,99 | 0.42-9.43 | 0.384 |
| Tiredness | 5.54 | 3.81-7.29 | 52.00 | 40.69-63.31 | 1,36 | 0.81-2.27 | 0.238 |
| Mild | - | - | - | - | 1,00 | 0.52-1.89 | 0.999 |
| Moderate | - | - | - | - | 2,18 | 0.86-5.49 | 0.099 |
| Severe | - | - | - | - | 2,17 | 0.88-5.31 | 0.089 |

<sup>1</sup> The model to investigate the severity did not converge, even grouping the severity in two categories.
<sup>2</sup> Incidence rate per 100 person/day is the number of a new reports per unit of person-time at risk.
<sup>3</sup> Cumulative incidence is the proportion of new reports among an initially disease-free population.
<sup>4</sup> Mixed logistic model with subject and school identifiers as random intercepts.

## Discussion

This study confirms the potential of a syndromic surveillance among children aged 6-11 years, as a complementary strategy to support the public health actions applying analytical models that improve their potential in providing timely and systematized information about the student’s health status, symptoms and their association with school absences.

We found significantly and positively association between school absences and common cold syndrome represented by the most common symptoms reported such as sore throat, sneezing and cough. Despite being one of the most frequently reported symptoms, tiredness was not associated with school absence, and considering that the questionnaire was applied at the end of the 21-22 academic year, this result was expected.

In this pilot study, we collected data on absences, with the support of the teacher, and on self-reported symptoms with primary school students, who answered a very simple questionnaire by themselves. Like us, other studies have also adapted tools to collect data from children, considering that they generally do not communicate their health status very well. [6,14].

The early key of this study was scaling the self-application of the survey for students of younger age groups, ensure their participation without increasing the burden of attributions of teachers and health services that have already been largely impacted by the SARS-CoV-2 pandemic. In the CSSNC project, we had already tested a self-administrated questionnaire for students over 16 years and antigen self-tests for children over 9 years, obtaining good results [16,18].

The global concern about public health emergencies demands the improvement of the strategies to monitoring the health status, risk factors and social determinants in populations at risk. It was possible, through this study, to observe a relationship between symptoms on a severity scale and school absence. We understand that the analytical model allowed us to find and explore trends of clustering of symptoms and association with the established outcome, even with a few days of monitoring, that is, it was possible to capture events within restricted periods of time. The integration of sentinel school surveillance with routine surveillance by collecting data on respiratory symptoms and school absence can help to better understand the impact of ILI in this population and to detect the occurrence of events with greater sensitivity.

The nonspecific approach of the syndromic surveillance could bring more sensibility to detecting a wider range of events and situations [19], being useful to adapt tools, promote the best approach, use and allocation of resources and ensure the best monitoring model [2,20,21]. Moreover, school absences have been already used as a tool for alerting public health events caused by influenza-like events.[6,11,13,22–25]

Considering the few number of students participants and that the exposures may occur elsewhere, we cannot extrapolate these results to the community overall. Considering the short study period, we cannot detect any seasonal variation. The strategy of syndromic surveillance must be active during a larger period, until it is possible to establish alert limits and the specific forecast models for the target’s diseases, real-time data based, for the school-age population. We were also unable to determine the health status in approximately half of the absences that were classified as by unknown reasons.

## Final considerations

This article presents the potential of monitoring school absences and syndromic surveillance in a school sentinel population during the COVID-19 pandemic in Catalonia. The pilot carried out with schoolchildren shows the implementation of a strategy adapted for primary school students, with the aim of monitoring their health status in addition to contextualizing school absence by associating it with the occurrence of symptoms among the participants. In addition, the analytical plan focused on exploring the occurrence of syndromes, constructed from the grouping of these data, enabling its application for the prediction of health events, offering either an opportunity for quick action, or simply for monitoring and understanding the students’ health status.

## Data Availability

All data produced in the present work are contained in the manuscript. Complementary anonymized databases and their corresponding codebooks can be available through formal proposal to the CEEISCAT via. Due to legal restrictions in relation to the Personal Information Protection Act, personal or spatial data that allow identified any participant, including the name of the school, which was used as an adjustment factor in the analysis, cannot be made publicly available.

## Acknowledgments

The authors would like to thank the Health Department and Department of Education of the Government of the Catalonia (Spain), the former D*irecció General de Recerca i Innovació en Salut* (DGRIS) and *Institut Català de la Salut* (ICS) that made this project possible, and all the health care professionals acting as a COVID-19 pandemic health taskforce in Catalonia. Also, we would like to specially thank the effort and dedication all the sentinel schools’ staff, students and families who participated in the project.

## COVID-19 Sentinel School Network Study Group of Catalonia (CSSNC)

### Principal investigators

Jordi Casabona [Centre d’Estudis Epidemiològics sobre les Infeccions de Transmissió Sexual i Sida de Catalunya (CEEISCAT)-CIBERESP], Josep Basora (Institut Universitari d’Investigació en Atenció Primària (IDIAP Jordi Gol).

### Project manager

Anna Bordas (CEEISCAT).

### Technical committee

Jordi Casabona (CEEISCAT), Jordi Sunyer (ISGlobal), Antoni Soriano (Hospital Universitari Vall d’Hebron), Rosina Malagrida (Living lab for Health, IRSICaixa)

### as Work Package coordinators

Cinta Folch, Pol Romano, Esteve Muntada, Anna Bordas, Fabiana Ganem, Andreu Colom i Jordi Casabona (CEEISCAT), Mireia Gascón, Maria Subirana, Pau Majo, Jordi Sunyer (ISGlobal), Rosina Malagrida, Laia Vives (Living lab for Health), Antoni Soriano, Pere Soler-Palacín (Hospital Universitari Vall d’Hebron).

### Microbiology laboratories

Tomàs Pumarola, Andrés Antón, Cristina Andrés, Juliana Esperalba, Albert Blanco (Hospital Universitari Vall d’Hebron), Ignacio Blanco, Pere-Joan Cardona, Maria Victoria González, Gema Fernández, Cristina Esteban (Hospital Universitari Germans Trias i Pujol)

### Data Management and statistical analysis

Yesika Díaz, Lucia Alonso, Jordi Aceiton, Marcos Montoro (CEEISCAT).

### Data Protection Officer and Technical Support

Esteve Muntada (CEEISCAT).

### Communication manager

Pol Romano, Cristina Sánchez (CEEISCAT).

### Field team

Maria Subirana, Pau Majo (ISGlobal), Jessica Fernández, Laia Vives (Living Lab for Health, IRSICaixa), Andreu Colom, Isabel Martínez, Marina Herrero, Alba García, Juan Rus, Paula Ribas, Alba Blanco (CEEISCAT).

### Universitat Autònoma de Barcelona team

Juan Leyva, Mariela Patricia Aguayo, David Giménez i Carolina Watson.

### Community Paediatricians

Esperança Macià i Silvia Burgaya (CAP Manlleu), Mª Teresa Riera-Bosch, Elisabet Sola (EAP Vic Nord), Lidia Aulet, Maria Mendoza, Lidia Busquets (EAP Vic Sud), Xavier Perramon, Júlia Sebastià (EAP Eixample Dret), Ana Moreno (Cap Ripollet), Xavier Duran, Belen Pérez (EAP Can Gibert del Pla), Anna Gatell (Equip Territorial de Pediatria Alt Penedès), Maria Coma (Hospital Universitari Joan XXIII).

### Ministry of Health

Carmen Cabezas (Secretaria de Salut Pública de Catalunya, Departament de Salut), Ariadna Mas (Direcció Assistencial Atenció Primària, Institut Català de la Salut), Maria Antònia Llopis (Coordinació dels laboratoris de l’Institut Català de la Salut), Josep Vidal (Gerència Territorial de la Catalunya Central, Institut Català de la Salut), Sandra Pequeño and Jacobo Mendioroz (Subdirecció general de Vigilància i Resposta a Emergències de l’Agència de Salut Pública de Catalunya, Departament de Salut), Robert Fabregat (former Direcció Gral Recerca i Innovació, Departament de Salut).

### Department of Education

Josep Gonzàlez-Cambray (Conseller d’Educació), Núria Mora (Secretaria de Transformació Educativa), Maria Neus Fornells (former Gabinet del Conseller), Rut Ribas (former Direcció general de l’alumnat).

## Data sharing

All data produced in the present work are contained in the manuscript. Complementary anonymized databases and their corresponding codebooks can be available through formal proposal to the CEEISCAT via. Due to legal restrictions in relation to the “Personal Information Protection Act,” personal or spatial data that allow identified any participant, including the name of the school, which was used as an adjustment factor in the analysis, cannot be made publicly available.

## Conflict of interest

The authors declares that the research was conducted in the absence of any commercial or financial relationships that could be construed as a potential conflict of interest.

## Author contributions

FG, CF, AB, ACC, AS and JC conceived and designed the study. ACC were responsible for the data collection; FG and LA performed the statistical and epidemiological analysis. FG wrote the first draft of the manuscript. All authors drafted the manuscript for important intellectual content, contributed to revision of the final version of the manuscript, approved the final version submitted, and agree to be accountable for all aspects of the work in ensuring that questions related to the accuracy or integrity of any part of the work are appropriately investigated and resolved. JC acts as guarantor. The corresponding author attests that all listed authors meet authorship criteria and that no others meeting the criteria have been omitted.

## Funding

This work was supported by the Health Department of the Government of Catalonia with no grant number. The funders had no role in study design, data collection and analysis, decision to publish, or preparation of the manuscript. FG, AB, LA and ACC received a salary from the abovementioned funder.

## Transparency statement

The corresponding author, Cinta Folch, on behalf of the rest of the signatories, guarantees the accuracy, transparency and honesty of the data and information contained in the study; that no relevant information has been omitted; and that all discrepancies between authors/authors have been adequately resolved and described.

## Notes

### Competing Interest Statement

The authors have declared no competing interest.

